# Prediction of Sepsis Mortality in ICU Patients Using Machine Learning Methods

**DOI:** 10.1101/2024.03.14.24304184

**Authors:** Jiayi Gao, Yuying Lu, Negin Ashrafi, Ian Domingo, Kamiar Alaei, Maryam Pishgar

## Abstract

**Problem:** Sepsis, a life-threatening condition, accounts for the deaths of millions of people worldwide. Accurate prediction of sepsis outcomes is crucial for effective treatment and management. Previous studies have utilized machine learning for prognosis, but have limitations in feature sets and model interpretability.

**Aim:** This study aims to develop a machine learning model that enhances prediction accuracy for sepsis outcomes using a reduced set of features, thereby addressing the limitations of previous studies and enhancing model interpretability.

**Methods:** This study analyzes intensive care patient outcomes using the MIMIC-IV database, focusing on adult sepsis cases. Employing the latest data extraction tools, such as Google Big- Query, and following stringent selection criteria, we selected 38 features in this study. This selection is also informed by a comprehensive literature review and clinical expertise. Data preprocessing included handling missing values, regrouping categorical variables, and using the Synthetic Minority Over-sampling Technique (SMOTE) to balance the data. We evaluated several machine learning models: Decision Trees, Gradient Boosting, XGBoost, LightGBM, Multilayer Perceptrons (MLP), Support Vector Machines (SVM), and Random Forest. The Sequential Halving and Classification (SHAC) algorithm was used for hyperparameter tuning, and both train-test split and cross-validation methodologies were employed for performance and computational efficiency.

**Results:** The Random Forest model was the most effective, achieving an area under the receiver operating characteristic curve (AUROC) of 0.94 with a confidence interval of ±0.01. This significantly outperformed other models and set a new benchmark in the literature. The model also provided detailed insights into the importance of various clinical features, with the Sequential Organ Failure Assessment (SOFA) score and average urine output being highly predictive. SHAP (Shapley Additive Explanations) analysis further enhanced the model’s interpretability, offering a clearer understanding of feature impacts.

**Conclusion:** This study demonstrates significant improvements in predicting sepsis outcomes using a Random Forest model, supported by advanced machine learning techniques and thorough data preprocessing. Our approach provided detailed insights into the key clinical features impacting sepsis mortality, making the model both highly accurate and interpretable. By enhancing the model’s practical utility in clinical settings, we offer a valuable tool for healthcare professionals to make data-driven decisions, ultimately aiming to minimize sepsis-induced fatalities.

## 1 Background

Sepsis can cause the failure of one or more organ systems, which is a life-threatening condition that occurs unpredictably and can progress rapidly [1–5]. By 2017, Sepsis accounted for nearly 20% of all global deaths; more specifically, there were 11 million sepsis-related deaths in total 48.9 million sepsis cases [6]. Among those, 1.7 million adults develop sepsis each year in the United States, which causes around 270,000 deaths [7]. In a 2020 study, Suveges and other examine [8] analyzed 110,204 hospital admissions, revealing a direct correlation between the length of hospital stay and survival, with an average stay of 10 days indicating a decreased likelihood of survival. Given the severity of the illness, it is crucial to find the possible factors that contribute to the mortality of sepsis [9–11].

Traditionally, various scoring systems (i.e. SOFA score) were used to predict in-hospital mortality for critically ill patients with sepsis [12–15]. Such systems, while effective, are often limited in the range of features they examine [16–18]. For example, these scoring systems typically focus on a narrow set of clinical parameters, which might not capture the full complexity of sepsis. This limitation can lead to incomplete assessments of a patient’s condition and subsequently, less accurate predictions. Other studies, such as retrospective analysis, are also popular methods for evaluating relationships between a specific feature and mortality. For instance, Bi’s study [19] demonstrates a correlation between PaO2/FiO2 levels and 28-day mortality, more specifically, on a 200mg threshold. However, even though accurate, these studies are less effective and can only examine one pair of relationship at a time. This approach does not account for the multifactorial nature of sepsis, where multiple physiological and biochemical parameters interact in complex ways. As a result, these studies often miss critical interactions between features that could improve the predictive accuracy of sepsis outcomes. The inability to integrate and analyze multiple features simultaneously poses a significant barrier to developing more comprehensive and precise predictive models for sepsis. Furthermore, the reliance on retrospective data means these models are often not adaptive to the dynamic and rapidly changing clinical status of sepsis patients, further limiting their real-time applicability and effectiveness.

To overcome the limitations of traditional methods, recent studies have pivoted towards Machine Learning (ML) and Deep Learning (DL) approaches [20–30]. In Bao’s study [6], they presented the efficacy of the Light GBM algorithm in predicting sepsis patient mortality, suggesting its integration into clinical tools. Similarly, Shifang et al. [31] highlighted the potential of Artificial Neural Networks (ANN) in the early detection of high-risk patients. Moreover, machine learning methods are increasingly being employed across a broad spectrum of medical-related topics, demonstrating their versatility and efficacy. [32–40], However, even though these previous works introduced advanced analytical methods, we found that they utilized a significant number of features and did not achieve satisfying results.These models often lacked comprehensive feature selection strategies and advanced data preprocessing techniques, which limited their accuracy and practicality in clinical settings. Additionally, the use of numerous features complicated the models, leading to overfitting and inefficiency, making them less suitable for real-time application. In the following, we will list the main contributions of our work:

- Advanced data preprocessing techniques were employed to address missing or duplicate values and to regroup categorical variables, significantly enhancing data quality and model performance.
- A thorough review of academic literature and recommendations from clinical experts guided our feature selection process, leading to more accurate predictions using a smaller, more relevant feature set.
- The use of SHAP (SHapley Additive exPlanations) analysis improved the interpretability of our model’s predictive outcomes, providing granular insights into the factors affecting sepsis mortality.
- The Synthetic Minority Over-sampling Technique (SMOTE) was used to address data imbalance, significantly improving the robustness of our model.
- Our proposed model, particularly the Random Forest model, achieved an AUROC of 0.94 with a narrower confidence interval, representing a 6.3% improvement compared to the best existing study.

This paper sets a new benchmark in the field, significantly improving model accuracy and efficiency, and making our model a practical tool for healthcare professionals. The use of machine learning methods in medicine provides an immediate and accurate second opinion, serving as an alternate source of confirmation for medical professionals. Mortality predictions derived from these models are valuable assets for resource management in hospitals, allowing for the refactoring of resources to prioritize patients in more desperate conditions. Additionally, predictive models facilitate more efficient use of healthcare services by enabling urgent treatment for patients at greater risk of death, ultimately helping to save more lives. These advancements enhance clinical decision-making and improve patient outcomes.

The rest of the paper is organized as follows: Section 2 Methods describes the data source and inclusion criteria, feature selection and data preprocessing, modeling, statistical analysis between cohorts, and variables impacts. Section 3 Results presents the cohort characteristics, evaluation metrics, and Shapley value analysis. Section 4 Discussion interprets the findings and their significance. Section 5 Limitations addresses the study’s constraints and potential weaknesses. Section 6 Future Work suggests directions for enhancing predictive capabilities and research extensions. Finally, Section 7 Conclusion summarizes the key contributions and clinical impact of the study.

## 2 Methods

### 2.1 Data Source and Inclusion Criteria

The data for this study were sourced from the Medical Information Mart for Intensive Care IV (MIMIC-IV), an authoritative and comprehensive database [41]. The database contains health records of the Beth Israel Deaconess Medical Center from 2008 to 2019 and includes over 40,000 unique patients from critical care units. The admission information was recorded into various tables, such as demographics, lab results, and ICU information. Compared to its predecessor, MIMIC-III, this dataset contains updated patient information and extends the scope of data captured, thus offering a more current view of patient care. The utilization of MIMIC-IV for our study ensures that our analysis is grounded in the latest available data, facilitating a more accurate and relevant exploration into the factors affecting patient outcomes in intensive care settings.

To narrow down the target patients, we applied the following criteria. These criteria stipulated that only adult patients (aged 18 and above) with a minimum intensive care unit (ICU) stay of over 24 hours were considered to guarantee ample data for a thorough analysis. Furthermore, the study targeted patients diagnosed with sepsis based on the Third International Consensus Definitions for Sepsis and Septic Shock (Sepsis-3), with a Sequential Organ Failure Assessment (SOFA) score of 2 or higher and a suspected infection as recorded in the MIMIC-IV database. This study implements BigQuery as the data extraction tool to select the target patients from the dataset.

### 2.2 Feature Selection and Data Preprocessing

The feature selection process was informed by a thorough review of academic literature and guided by recommendations from a clinical expertise. The selection methodology took two key considerations into account: (1) the recurrence of specific features across multiple studies, signaling their widespread recognition in critical care, and (2) the acknowledgment of certain features in prior individual studies as vital for mortality prediction. This selection was based on their prevalence in existing literature, clinical importance, and statistical validation to ensure their relevance and predictive power.

The final dataset contains 38 distinct features, including demographic information, antibiotic usage, patient medical history, and various laboratory results. Variables such as the Sequential Organ Failure Assessment (SOFA) score, average urine output, minimum and maximum glucose levels, sodium levels, heart rate, systolic and diastolic blood pressures (SBP and DBP), respiratory rate, oxygen saturation (SPO2), and albumin levels. These features were selected due to their frequent appearances in related studies, emphasizing their predictive value for patient outcomes. By integrating these variables, the dataset provides a robust foundation for developing predictive models, aiming to enhance the accuracy of mortality and prognosis estimations in critical care settings.

Further improving the dataset, we set a threshold for the PaO2/FiO2 ratio of 200 [17]. Additionally, based on the recommendation of a clinical expertise, the coma score was incorporated, categorizing patients with scores above 8 as in a coma. Following the feature selection process, the dataset was narrowed down to 6,401 admission records. A detailed list of features, along with the categories they fall into, can be found in the Table 1 provided below.

**Table 1:** Detailed Overview of Feature Information.

| Feature Type | Feature Name | Feature Type | Feature Name |
| --- | --- | --- | --- |
| Admission Information | los_icu | Demographics | Age |
| Lab Results | SOFA_score | Comorbidities | diabetes_without_cc |
|  | avg_urineoutput |  | diabetes_with_cc |
|  | temperature_min |  | severe_liver_disease |
|  | temperature_max |  | aids |
|  | temperature_avg |  | renal_disease |
|  | heart_rate_min | Medications |  |
|  | heart_rate_max |  |  |
|  | heart_rate_mean |  | antibiotic_Carbapenem |
|  | resp_rate_min |  | antibiotic_Aminoglycoside |
|  | resp_rate_max |  | antibiotic_Glycopeptide |
|  | resp_rate_mean |  | antibiotic_Oxazolidinone |
|  | spo2_min |  | antibiotic_Penicillin |
|  | spo2_max |  | antibiotic_Sulfonamide |
|  | spo2_mean |  | antibiotic_Tetracycline |
|  | hospital_expire_flag |  |  |

The dataset’s cleaning was approached with the following steps: (1) addressing null values and duplicates in both numerical and categorical data; (2) grouping the existing categorical variables (race and antibiotics) into new features to facilitate future encoding processes. Specifically, races were separated and summarized into four groups: Black or African American, Hispanic or Latinx, White, and Other Races. For the antibiotics, the existing 25 categories were regrouped into seven different groups based on their chemical structure, mechanism of action, spectrum of activity, side effects, and toxicity. These groups are Aminoglycosides, Carbapenems, Glycopeptides, Oxazolidinones, Penicillins, Sulfonamides, and Tetracyclines.

Upon review, the training data is imbalanced, which is a common issue in healthcare datasets. Unlike the cluster centroids method used in existing literature, the Synthetic Minority Over- sampling Technique(SMOTE) method was introduced to address this data imbalance issue by oversampling [42]. SMOTE method helps raise our data points for the minority class, which increases the likelihood that models will generalize well to new, unseen data and reduces the risk of overfitting. After applying the SMOTE method, the data points expanded from 6,401 to 7,304. By doing so, SMOTE helps balance the dataset, which is crucial for training models that generalize well to new, unseen data and reduces the risk of overfitting. This method ensures that our predictive models are more robust and reliable. Below is Figure 1, which illustrates the workflow for data preprocessing.

**Figure 1:**
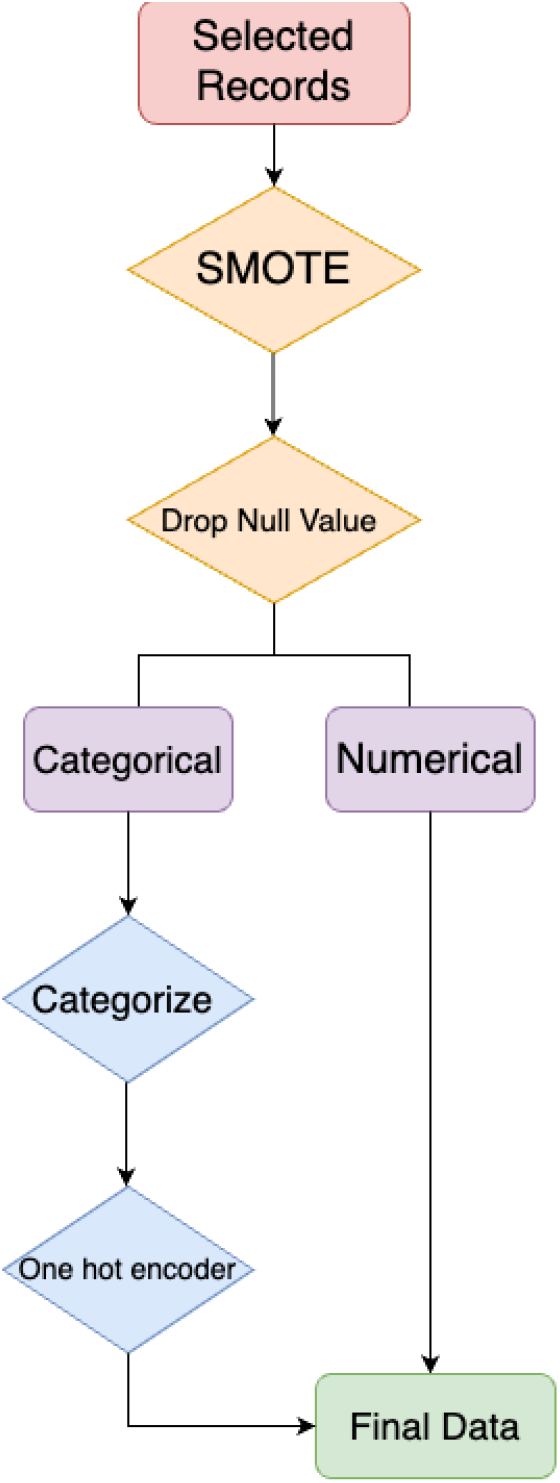
Work flow of data preprocessing. The process begins with selected records (red), followed by initial preprocessing steps like SMOTE and dropping null values (orange). Data is then divided into categorical and numerical types for further processing (purple). Categorical data is categorized and encoded (blue). Finally, the processed data is ready for model fitting (green).

### 2.3 Modeling

The final dataset comprises 53 columns and 7,304 data points and has achieved balance after the application of SMOTE. To thoroughly evaluate the performance of various machine learning classification models, we utilized two methodologies: (1) train-test split; (2) 5-fold cross-validation and hyper-parameter tuning. More specifically, the Sequential Halving and Classification (SHAC) algorithm, proposed by Kumar et al. [43], was adopted as a more efficient alternative to exhaustive grid search for hyperparameter tuning and preventing overfitting. We then fed the resulted dataset to the following models: tree-based models such as Decision Trees [44], ensemble methods like Gradient Boosting [45], Extra Gradient Boosting (XGBoost) [46], Light Gradient Boosting Machine (LightGBM) [47], neural networks with a focus on Multilayer Perceptrons (MLP) [48], margin-based models including Support Vector Machines (SVM) [49], and bagging models, notably Random Forest [50].

To determine the proposed model, we meticulously evaluated three key factors: firstly, the Area Under the Receiver Operating Characteristic (AUROC) scores to assess accuracy; secondly, sensitivity to variance, serving as a gauge for the model’s robustness; and thirdly, the overall consistency to ensure reliability across different datasets. Consequently, this evaluation framework showed that the Random Forest model outperformed other models using both the train-test split and cross-validation methodologies. This choice was driven by the model’s superior AUROC scores, affirming its effectiveness in prediction and its potential for handling new data. Our selection process highlights the significance of utilizing a structured evaluation to identify a model that not only shows high performance but also maintains robustness and consistency under various conditions. Figure 2 below shows the workflow of our methodology.

**Figure 2:**
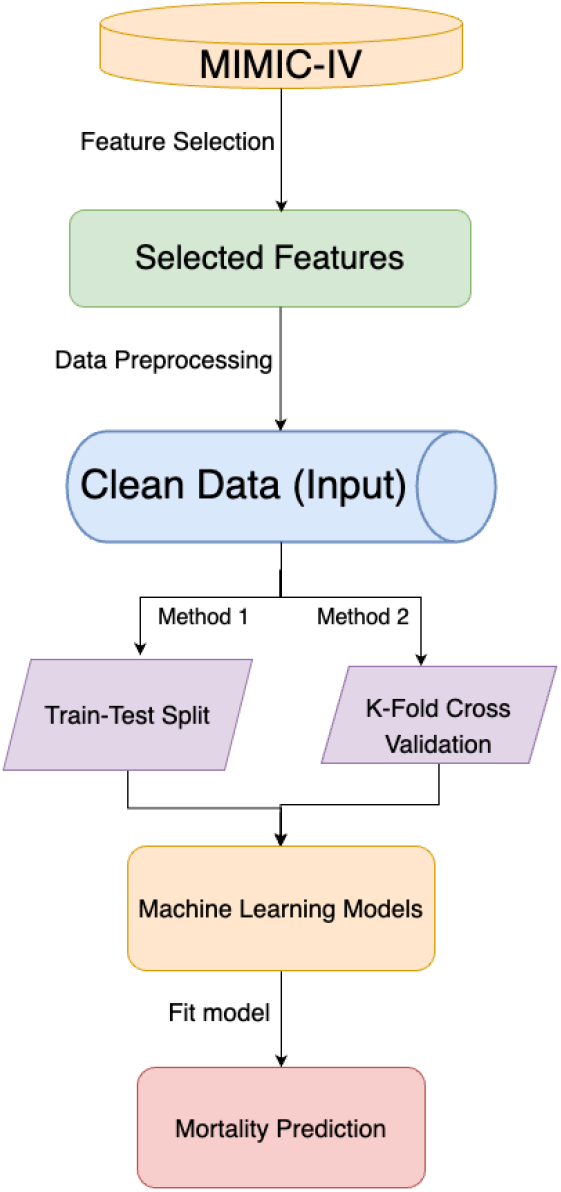
Overview of the Methodology. The process starts with extracting target patients from MIMIC-IV, followed by feature selection and data preprocessing (green). The input data (blue) is split for model fitting (purple). The optimal model (yellow) is then used for predictions (red).

### 2.4 Statistic analysis between cohorts

Statistical analyses, such as the chi-square test and two-sided t-test, were performed to compare the measurements of variables in the train and test cohorts. More specifically, the comparison for categorical features was conducted using the chi-square test, while for numerical features, the two-sided t-test was employed. These model developments and statistical tests were conducted in Python version 3.6.

### 2.5 Variables Impacts

Shapley value analysis [51] was performed on the test set to determine the influence of each variable on the predictions of our proposed model and to identify the variable most closely linked to mortality. The Shapley values illustrated the average impact of each variable on the results within various groups [52]. In comparison to traditional feature importance measures, such as those derived from Random Forest, Shapley values offer a more comprehensive understanding of the impact of each feature on model predictions. While Random Forest feature importance typically relies on metrics like Gini impurity or information gain, SHAP values consider the entire space of possible feature combinations and allocate contributions fairly among features [53]. The key distinction lies in the interpretability of Shapley values on an instance level, allowing us to understand the specific influence of each feature for a given prediction. This level of granularity is especially valuable when dealing with complex models and real-world datasets.

## 3 Results

### 3.1 Cohort Characteristics Model Completion

Following the discussion of feature selection and data preprocessing, the 7304 data points were used to train the model. The number of data points was determined by setting thresholds for LnPaO2/FiO2 at 200 mg, based on a literature review of Bi’s study, which highlighted LnPaO2/FiO2 as an important factor influencing sepsis mortality. Additionally, the SMOTE method was employed to balance the dataset [19]. These data points were split into train and test cohorts randomly with a ratio of 0.8 to 0.2. The model with the best AUROC was chosen to further evaluate the test set.

In terms of ICU stay, the average length in the training cohort was 6.974 days, compared to 6.977 days in the testing cohort. Given that the p-value from the two-sided t-test is 0.989, surpassing our predetermined alpha threshold of 0.05, this indicates no significant difference in ICU stay length between the two cohorts. Regarding age, the training set had an average age of 65.160 years, slightly lower than the testing set’s average of 66.039 years. However, the p-value from the two-sided t-test here is 0.086, suggesting no statistically significant age difference between the cohorts. All other features, except for urine output, show no statistical significance, which indicates a notable difference in average urine output between the training and testing cohorts. Table 2 below presents the statistical results of train and test cohorts for both numerical and categorical variables, where numerical feature values represent the average measurements, and categorical feature values represent the percentage of individuals in each category.

**Table 2:** Statistical Analysis Results of Train and Test Cohorts for Numerical and Categorical Variables.

| Feature | Train | Test | P-value |
| --- | --- | --- | --- |
| *Admission Information* |  |  |  |
| los_icu | 6.974 | 6.977 | 0.989 |
| *Demographics* |  |  |  |
| Age | 65.160 | 66.039 | 0.086 |
| gender_F | 42.188 | 41.842 | 0.848 |
| *Lab Results* |  |  |  |
| SOFA_score | 4.317 | 4.393 | 0.305 |
| avg_urineoutput | 160.971 | 153.108 | 0.007 |
| temperature_min | 36.663 | 36.632 | 0.478 |
| temperature_max | 37.149 | 37.145 | 0.920 |
| temperature_avg | 36.906 | 36.888 | 0.652 |
| glucose_min | 110.363 | 111.451 | 0.414 |
| glucose_max | 211.147 | 214.524 | 0.453 |
| glucose_average | 154.669 | 156.739 | 0.332 |
| sodium_min | 135.376 | 135.385 | 0.961 |
| sodium_max | 141.558 | 141.768 | 0.222 |
| sodium_average | 138.499 | 138.595 | 0.521 |
| heart_rate_min | 70.250 | 69.694 | 0.271 |
| heart_rate_max | 114.752 | 114.909 | 0.819 |
| heart_rate_mean | 89.384 | 89.530 | 0.763 |
| sbp_min | 81.941 | 81.595 | 0.489 |
| sbp_max | 154.473 | 153.932 | 0.475 |
| sbp_mean | 114.334 | 114.630 | 0.495 |
| dbp_min | 41.534 | 41.300 | 0.490 |
| dbp_max | 93.673 | 93.545 | 0.849 |
| dbp_mean | 61.570 | 61.466 | 0.725 |
| resp_rate_min | 11.932 | 11.804 | 0.307 |
| resp_rate_max | 31.218 | 31.304 | 0.706 |
| resp_rate_mean | 20.510 | 20.529 | 0.881 |
| spo2_min | 89.180 | 89.023 | 0.586 |
| spo2_max | 99.732 | 99.755 | 0.362 |
| spo2_mean | 96.887 | 96.868 | 0.801 |
| hospital_expire_flag | 28.242 | 30.445 | 0.127 |
| *Comorbidities* |  |  |  |
| diabetes_without_cc | 31.641 | 33.880 | 0.133 |
| diabetes_with_cc | 10.117 | 9.446 | 0.506 |
| severe_liver_disease | 10.449 | 9.758 | 0.499 |
| aids | 1.035 | 0.937 | 0.874 |
| renal_disease | 26.738 | 27.244 | 0.741 |
| *Medications* |  |  |  |
| antibiotic_Carbapenem | 15.664 | 14.910 | 0.533 |
| antibiotic_Aminoglycoside | 7.168 | 7.962 | 0.360 |
| antibiotic_Glycopeptide | 65.039 | 66.042 | 0.521 |
| antibiotic_Oxazolidinone | 0.195 | 0.156 | 1.000 |
| antibiotic_Penicillin | 0.000 | 0.078 | 0.453 |
| antibiotic_Sulfonamide | 0.723 | 1.093 | 0.247 |

Table 2 – Continued from previous page
| Feature | Train | Test | P-value |
| --- | --- | --- | --- |
| antibiotic_Tetracycline | 0.000 | 0.078 | 0.453 |

### 3.2 Evaluation metrics proposed and baseline models performance

Figure 3 below illustrates the ROC curves for each model along with their corresponding AUC values. Notably, except for the Decision Tree’s curve, other curves exhibit remarkably smooth shapes. Additionally, it was noted that more complex models, particularly those based on ensemble learning techniques like XGB and LGBM, outperformed simpler base models. For instance, both XGB and LGBM achieved impressive AUC values of 0.92, significantly higher than the 0.75 attained by simpler models such as Decision Trees and SVM.

**Figure 3:**
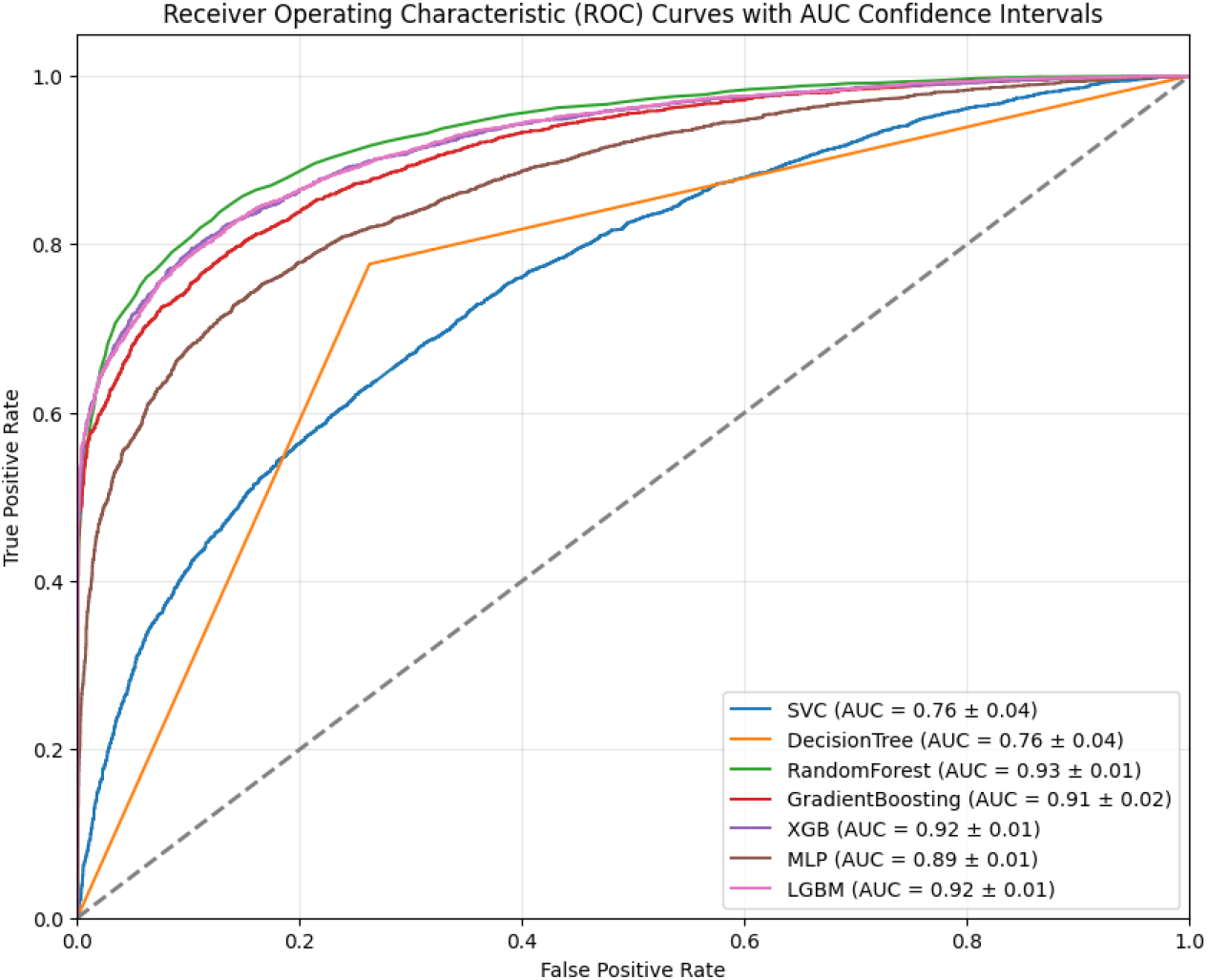
The figure illustrates the ROC AUC scores and the confidence intervals of mortality prediction machine learning models. It shows that the model with the highest AUROC score and the most stable confidence interval is RandomForest.

The table (Table 3) below presents the detailed results, including other evaluations such as sensitivity, specificity, and F1 score. For both train test split and cross-validation, random forest demonstrated the best result. Hence, Random Forest (RF) is the proposed model in this paper, which achieved a 0.94 AUC score with a 0.01 confidence interval representing a 6.3% improvement compared to the best existing study. It is important to note that, although we typically expect cross-validation to provide a more robust estimate of model performance, in this particular study, the train-test split yielded slightly better results by chance. By using a random seed of 42 to split our dataset into training and test sets, we achieved an AUROC of 0.93, which was 1% higher than the cross-validation results. Both methods demonstrated high AUROC scores and narrow confidence intervals, indicating the stability and robustness of the dataset and features selected. This consistency across different data partitioning methods validates the reliability of our chosen approach and ensures that the model’s performance is not significantly affected by the method of data partitioning.

**Table 3:** Metrics for Baseline Models on Test Set and Random Forest on Different Sets.

| Model | AUROC | Precision | Sensitivity | Accuracy | F-score |
| --- | --- | --- | --- | --- | --- |
| Decision Tree | 0.7329 | 0.7219 | 0.7475 | 0.7327 | 0.7345 |
| SVC | 0.7277 | 0.6772 | 0.6179 | 0.6654 | 0.6462 |
| GradientBoosting | 0.906 | 0.8469 | 0.7597 | 0.8133 | 0.8009 |
| XGB | 0.9192 | 0.8491 | 0.8228 | 0.8401 | 0.8358 |
| MLP | 0.8763 | 0.7495 | 0.835 | 0.7804 | 0.7899 |
| LGBM | 0.9185 | 0.8521 | 0.7973 | 0.8313 | 0.8238 |
| RF (Test Set) | 0.9388 | 0.876 | 0.8372 | 0.8631 | 0.857 |
| RF (Trainig Set) | 1.0000 | 1.0000 | 1.0000 | 1.0000 | 1.0000 |
| RF (Cross Validation) | 0.9293 | 0.8599 | 0.8312 | 0.8475 | 0.8453 |

### 3.3 Shapley Value analysis

SHAP values analysis was applied to evaluate the importance of the feature within the context of Random Forest. According to the results of the SHAP analysis, the coma score has the highest impact on mortality prediction, which indicates that higher coma scores tend to have a strong positive impact on the model’s prediction of mortality. In other words, as the coma score increases, the likelihood of mortality, as predicted by the model, also increases. Additionally, average urine output shows a notable impact. Lower average urine outputs are more influential in increasing the prediction of mortality compared to high urine outputs, suggesting that lower average urine outputs are associated with an increased prediction of mortality. Regarding the feature ‘gender Male’, since a high SHAP value correlates with a decrease in the model’s prediction of mortality and males are represented by one, it indicates that being male is associated with a lower risk of mortality compared to females. Figure 4 demonstrates the detailed result of the SHAP values analysis. The ‘Sum of 39 features’ in the SHAP plot represents the combined SHAP values of the remaining less influential features. This aggregation provides a holistic view of their cumulative impact on the model’s predictions, highlighting that while individually these features may not have a significant impact, together they can still influence the model’s outcomes.

**Figure 4:**
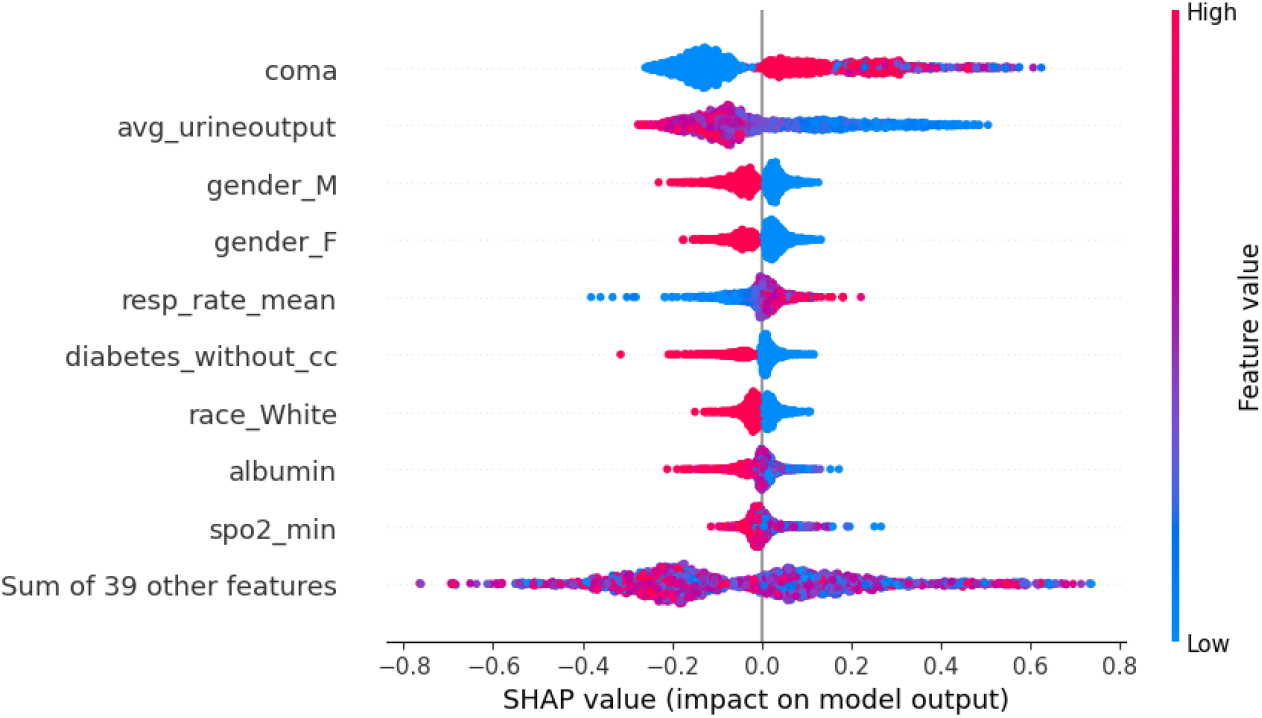
This figure illustrates the feature importance, impact direction on mortality prediction, and distribution of Shapley values for each feature. The figure indicates that coma, avg urineoutput, and gender M have the highest impact on the prediction. The dots on the right are mostly red, meaning that when the feature has a higher value, it will increase the probability of mortality.

After reviewing other articles about sepsis mortality rates, we found that there is some overlap between the top features. For example, the top five features in Bao’s study are glucose max, urine output, platelets max, age, and MBP max. This overlaps with our result on urine output, indicating that urine output is an important feature to be considered in real-life situations. These insights highlight key factors affecting mortality. Understanding that higher coma scores and lower average urine outputs significantly increase the risk of mortality, while being male is associated with a lower risk, can help in developing targeted interventions to mitigate mortality risks.

## 4 Discussion

In this study, supervised machine learning models were used to forecast mortality from sepsis over 24 hours of ICU admission. Our approach involved selecting a group of high-impact features, ensuring a concise and relevant model. To address the issue of dataset imbalance, we implemented the SMOTE, significantly bolstering the robustness and dependability of our predictive model. Furthermore, we employed SHAP analysis to identify and quantify the contribution of each feature to our model’s outcomes, thereby enhancing the interpretability of our predictions in a clinical context.

The best AUROC curve achieved was 0.94 +/- 0.01. This is approximately 6.3% higher than the best result in our literature review, which is 0.884. The narrower confidence interval indicates that our model is more stable. The higher AUROC means that the model is good at accurately predicting the mortality of patients by successfully discriminating between positive cases (hospital exp flag = 1, the patient died) and negative cases (hospital exp flag = 0, the patient does not die). The stability of the model is important for providing consistent prediction results and ensures more robustness against changes in input data, making the model’s outcomes more reliable. Furthermore, the study’s approach contributed to reducing sepsis-induced fatalities by providing personalized suggestions for each patient through model fitting. The model aids clinicians in making early identifications of sepsis, ensuring more attention is given to patients at higher risk of sepsis mortality. This early detection and targeted intervention enhance healthcare efficiency and effectiveness, ultimately helping to reduce sepsis-induced fatalities. Moreover, even though advanced analytical methods were applied in existing literature, they have obvious drawbacks. One such limitation is the wide confidence interval, which indicates the model’s performance might not be consistently high across different test sets. Another limitation is the employment of an excessive number of features. This overabundance could negatively impact the model’s predictive efficiency and interpretability, thus making it more difficult to use in clinical settings because of the increased complexity and risk of overfitting.

Our study has several advantages compared to previous studies. First, the SMOTE method helped deal with the data imbalance issue, which is one of the major reasons the model results improved. Another significant advantage of our methodology is the meticulous selection of features; with just 38 features—approximately half the average reported in the literature—we not only attained higher AUC scores but also achieved increased stability in our models. This deliberate minimization of features resulted in a 6.3% uplift in performance outcomes, alongside a narrower confidence interval, highlighting the efficacy and dependability of our approach. Furthermore, the use of advanced analytics provided valuable insights into key mortality factors, enhancing clinical decision-making and patient outcomes.

## 5 Limitation

Although significant improvements have been made in both features and results, leading to a more stable model, this study still has some limitations. Currently, the MIMIC-IV dataset is the only data source for mortality prediction, with no other dataset available for validation. Moreover, the complexity of machine learning algorithms can lead to difficulties in deciphering their decision-making pathways, posing a substantial obstacle for clinicians who require transparent and interpretable models. Last, as the fast development in the field of medicine, using historical datasets might not fully capture the latest clinical practices or treatments. Therefore, it is crucial to regularly update these datasets and incorporate new medical knowledge and technologies to ensure the models trained on them remain relevant and effective.

## 6 Future Work

For future studies, it would be advantageous to include additional datasets, such as the eICU Collaborative Research Database, to serve as validation sets. This approach would ensure the model’s robust performance across diverse patient data. Moreover, in addressing the model interpretability problem, our aim is to develop algorithms that not only predict with high accuracy but also provide explanations for their predictions. Furthermore, establishing a real-time data flow for immediate predictions of sepsis mortality is another objective. To enhance the efficiency of the study, future implementations might leverage data streamlining tools, such as Google Cloud Dataflow.

## 7 Conclusion

Our study has achieved significant advancements in predicting sepsis outcomes by utilizing advanced machine learning techniques and sophisticated data preprocessing methods. These methods include data grouping and effective solutions to data imbalance issues found in the MIMIC-IV database. Remarkably, our approach is characterized by its efficiency, relying on a limited number of features to generate highly accurate predictions, as indicated by a robust AUROC score and enhanced stability, which is reflected in a narrower confidence interval. As the number of variables decreased, the model became more stable compared to the results in the literature, which used many more features. Second, the AUROC for this study is higher compared to other sepsis mortality prediction papers. From a real-life perspective, fewer features are more interpretable, which can help doctors and clinicians focus on the features that are more related to sepsis mortality. For the critical task of interpreting feature importance, we have incorporated the SHAP analysis, known for its consistency and the ability to provide a detailed explanation that is comprehensible to audiences from varied backgrounds. The following new standards have been established: the incorporation of diverse data types, including laboratory, demographic, and electronic health record data, and the use of advanced feature engineering methods that combine literature review and clinical insights. From this study, it can be concluded that patients with higher coma scores, lower average urine outputs, and female gender are more likely to be threatened by sepsis mortality according to the model’s predictions. In the future, clinicians can use the advanced machine learning model as a tool to identify patients with features that make them more susceptible to sepsis mortality. This allows clinicians to take proactive measures to decrease the mortality rate.

Our study has set new standards for predicting sepsis mortality by incorporating comprehensive data perspectives, including laboratory data, demographic data, and electronic health records. We also implemented advanced feature engineering methods, such as feature comparison with existing literature and real-world case suggestions from clinicians, to ensure accuracy and reliability in our predictions.

Additionally, the findings of this study substantiate the effectiveness of machine learning models in prognosticating sepsis. First, machine learning models can provide personalized suggestions for each patient through model fitting. Second, the model can help clinicians with the early identification of sepsis and ensure more attention is given to patients who are more likely to be affected by sepsis mortality, thereby increasing healthcare efficiency. As clinicians use the predictive model, it can enhance the efficiency of early diagnosis, provide personalized treatment plans for different patients, and improve and support the decision-making process. The notable precision of these models, coupled with the reduced breadth of confidence intervals, corroborates their reliability in generating consistent predictions, an attribute that is highly valued in clinical settings. Although it is imprudent for medical professionals to depend solely on machine learning models for the diagnosis and prognosis of medical conditions, these computational tools can serve as an adjunct, facilitating the confirmation of diagnostic outcomes or prompting a reevaluation of a patient’s status.

Our research underscores the potential of machine learning in clinical decision-making and prognostication within critical care settings. By employing these innovative approaches, we are moving towards a future where data-driven insights have the power to not only predict but also prevent sepsis-induced fatalities. The integration of such predictive models into clinical workflows could revolutionize patient care, offering clinicians a valuable tool in their efforts to combat this life-threatening condition.

## Data Availability

https://github.com/yuyinglu2000/Sepsis-Mortality.git

https://physionet.org/content/mimiciv/2.2/

## 8 Declaration

### 8.1 Ethics approval and consent to participate

The data supporting the findings of this article is available in the Medical Information Mart for Intensive Care version IV (MIMIC-IV). This publicly accessible, de-identified database did not require informed consent or Institutional Review Board approval. All procedures were conducted in accordance with applicable guidelines and regulations.

### 8.2 Consent for publication

Not Applicable

### 8.3 Availability of data and materials

The raw dataset is available in the MIMIC-IV repository:https://physionet.org/content/mimiciv/2.2/; and https://github.com/yuyinglu2000/Sepsis-Mortality.git

### 8.6 Authors’ contributions

J.G., Y.L., N.A., and M.P. invovled in all aspect of the study. I.D. formatted the manuscript and wrote part of the Background section. K.A. offered expert clinical advice. All authors reviewed and approved the final manuscript.

## 8.7 Acknowledgement

Notably, Jiayi Gao and Yuying Lu have contributed equally to this work. We also extend our gratitude for the publication of MIMIC IV.

